# The impact of WHO’s Treat All guideline on disease progression among people enrolled in HIV care in Central Africa: an observational cohort data by target trial design with multistate modeling

**DOI:** 10.1101/2022.08.27.22279144

**Authors:** Jiaqi Zhu, Hongbin Zhang, Ellen Brazier, Olga Tymejczyk, Marcel Yotebieng, April D. Kimmel, Kathryn Anastos, Jonathan Ross, Donald R Hoover, Qiuhu Shi, Gad Murenzi, Dominique Nsonde, Anastase Dzudie, Patricia Lelo, Christella Twizere, Denis Nash

**Affiliations:** Institute for Implementation Science in Population Health, City University of New York, New York, New York, United States of America; Graduate School of Public Health and Health Policy, City University of New York, New York, New York, United States of America; Department of Biostatistics, College of Public Health, University of Kentucky, Lexington, United States of America; Division of General Internal Medicine, Department of Medicine, Albert Einstein College of Medicine, Bronx, New York, USA; Department of Health Behavior and Policy, Virginia Commonwealth University School of Medicine, Richmond, VA, USA; Department of Statistics and Institute for Health, Health Care Policy and Aging Research, Rutgers the State University of New Jersey, Piscataway, NJ, USA; Department of Epidemiology and Community Health, New York Medical College, Valhalla, NY, USA; Einstein-Rwanda Research and Capacity Building Program, Rwanda Military Hospital and Research for Development (RD Rwanda), Kigali, Rwanda; CTA Brazzaville, Brazzaville, Republic of Congo; Clinical Research Education, Networking and Consultancy, Yaounde, Cameroon; Pediatric Hospital Kalembe Lembe, Kinshasa, Democratic Republic of Congo; Centre National de Reference en Matiere de VIH/SIDA, Bujumbura, Burundi

**Author notes:** Corresponding authors: D. Nash. Alternative corresponding author: H. Zhang.

**Keywords:** Treat All, HIV disease progression, Multistate model, Target trial, HIV care

## Abstract

WHO’s Treat All guidelines, which eliminate eligibility thresholds for people living with HIV to receive antiretroviral therapy, have been implemented by most countries. However, the impact of Treat All on the process of HIV disease progression is unknown. We conducted a target trial to emulate a hypothetical RCT to evaluate the policy’s impact on HIV disease progression among people living with HIV. We included people enrolled in HIV care during 2013-2019 from the Central Africa International Epidemiology Databases to Evaluate AIDS. Multistate models inferred the transitional hazards of disease progression across the four WHO clinical stages (1: asymptomatic; 2: mild; 3: advanced; 4: severe) and death. We estimated hazard ratios (HR) between a cohort enrolling in HIV care after (n=4,607) and a cohort enrolling before (n=4,439) Treat All guideline implementation, with and without covariates adjustment. Treat All implementation was associated with decreased hazards of transition in most stage categories, with significant results from stage 1 to stage 2 (adjusted HR (aHR) 0.64, 95% CI 0.44-0.94) and from stage 1 to death (0.37, 0.17-0.81), and non-significant but low HR results from stage 2 to 3 (0.71, 0.50-1.01), from stage 2 to death (0.58, 0.18-1.80). Treat All implementation substantially reduced HIV disease progression.

**Main Point Summary:** We compared the HIV disease progression outcome between a pri- and post-Treat All periods, utilizing individual service delivery data from Central Africa International Epidemiology Databases to Evaluate AIDS. We concluded that Treat All implementation substantially reduced HIV disease progression.

## Introduction

In September 2015, the World Health Organization (WHO) recommended immediate ART initiation for people living with HIV (PLWH), regardless of CD4 cell count or disease stage^1^. Since then, WHO’s Treat All recommendations have been adopted and implemented around the globe. By 2018, 84% of low-and middle-income countries had adopted the Treat All policy ^2^. Meanwhile, an increasing number of studies have evaluated the impact of Treat All policies ^3^. For example, it was shown that Treat All (1) increases timely ART initiation ^4,5^, (2) leads to a higher rate of viral suppression ^6^, and (3) reduces the risk of mortality ^7,8^. However, little is known about the role of Treat All guideline implementation on the *process* of HIV disease progression. For example, we are interested in the following questions for people living with HIV.

- How does Treat All policy implementation impact the progression among the health conditions (normal/mild/advanced/severe)?
- Did Treat All implementation reduce hazards for each health condition to mortality?

In resource-poor settings, where CD4 and HIV viral load testing are limited ^9^, the WHO HIV/AIDS Clinical Staging System is an important proxy to assess and monitor HIVpatients’ disease status ^10^. There are four stages in the staging system: stage 1 (asymptomatic), stage 2 (mild symptoms), stage 3 (advanced symptoms), and stage 4 (severe symptoms, i.e., AIDS). The staging system is a valid and practical way for prognosis and guides clinical management of people living with HIV ^5,11-15^. In this study, we aimed to evaluate the impact of the Treat All policy adoption on HIV disease progression, as defined by WHO clinical staging, and mortality, using data from the Central Africa cohort of the International Epidemiology Databases to Evaluate AIDS consortium (CA-IeDEA).

Due to the observational nature of the CA-IeDEA data, we took a target trial ^16,17^ approach to emulate a hypothetical randomized controlled trial. In addition, we used the multistate modeling method^27,28^ to quantify the HIV disease progression and to estimate the effect of Treat All on the process, utilizing the individual-level data.

## Materials and Methods

### Data Source

IeDEA (https://www.iedea.org) is an international research consortium of HIV cohorts sponsored by the National Institutes of Health ^18-20^. This study used data from 21 local clinics in five countries comprising the Central Africa IeDEA (CA-IeDEA): Burundi, Cameroon, Democratic Republic of Congo, Republic of Congo, and Rwanda from 2013 to 2019. Clinics participating in the CA-IeDEA cohort contribute individual-level longitudinal data on patients’ clinic visits, laboratory measurements, medications, and clinical outcomes.

### The Target Trial

We adopted the principle of target trial design ^16,17^ to emulate a hypothetical randomized controlled trial to assess the effect of *treatment* (aka, adopting the WHO’s recommendations on ART initiation at the national level). For each country, we constructed two mutually exclusive cohorts: the Treat All Guideline (TAG) cohort, which included patients enrolled in care after the adoption of the Treat All policy in each country, and the Non-Universal Guideline (NUG) cohort, which included patients enrolled in care before each country’s country adopted Treat All. Figure 1 shows a schematic diagram of the study design for Rwanda. Specifically, we started with each country’s Treat All adoption date and created a +/-3-month buffer window around the adoption date to accommodate the possibility of early or late site-level adoption of the policy relative to the national policy date ^21^. The database closing date determined the length of follow-up for the TAG cohort. Using the length, the starting date for the NUG cohort was determined to ensure same length of follow-up between the two cohorts.

**Figure 1.**
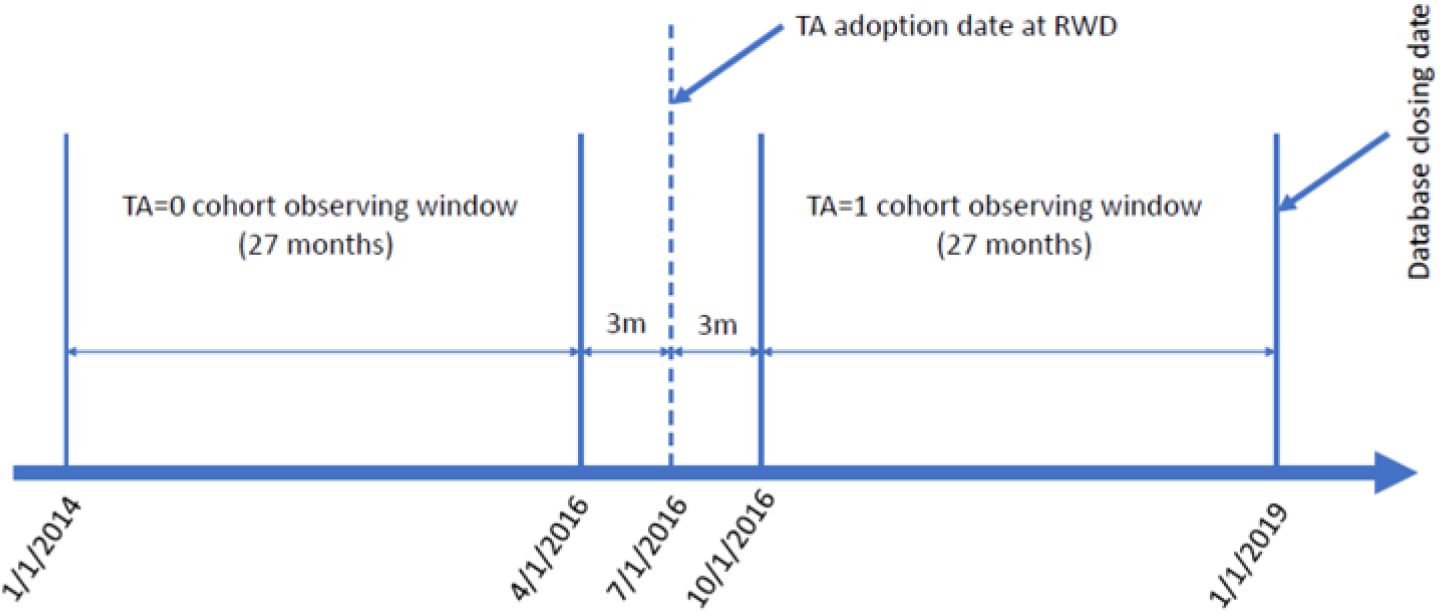
Schematic diagram of ‘cohorts’ construction under the target trial design for Rwanda.

#### Inclusion/Exclusion criteria

A patient was *eligible* for the trial if their enrollment date in HIV care was within the country’s respective TAG or NUG cohort window. We *excluded* patients who had no WHO stage at enrollment.

#### Intervention assignment

The TAG and NUG cohort constructed above emulated the *treatment* and *control* assignment for the hypothetical trial, without randomization, for the observational data. All patients in the TAG arm received Treat All policy intervention.

#### Outcomes

For each patient, we recorded their disease history from cohort entry, i.e., *time zero*, which was the HIV care enrollment date, to cohort exit, e.g., the end of cohort date or date of transfer out or death. We used *days from the enrollment* on all the entries and the corresponding state for the disease history recording. Here, the state refers to HIV clinical stage, according to the WHO Clinical Staging System (stage 1 to 4) and documented death (stage 5). Besides the five clinical states, we assigned two right censoring states and their corresponding time in our analysis, one at the time a patient transferred out and one at the end of cohort time. The observed measures of state status per timepoint constituted the event history data, i.e., the study’s survival *outcomes*.

### Analysis Strategy

We applied a Multistate Modeling (MSM) approach for the event history outcome (see below for the description of the MSM). The modeling was done through regression analyses, where the *treatment effects* were estimated as regression coefficients (log hazard ratio). For example, we included a binary covariate for Treat All policy adoption with a value of 1 for people in the TAG cohort and 0 for the NUG cohort. In a univariate model, we included the treatment variable only to assess the unadjusted effect of Treat All.

Without randomization, we expected a certain degree of imbalance between the two arms (TAG vs. NUG) on the outcome at baseline. Therefore, while the MSM enabled quantifying and comparing the underlying HIV disease process starting from time zero, we included other available measures at baseline in the models, emulating the *covariates adjustment* procedure in RCT. Specifically, in the adjusted analyses, we had confounders: age at enrollment, gender, season at enrollment, country, marital status, and CD4 count at baseline.

Our analysis was to emulate an *intention-to-treat* analytical approach, focusing on the overall effectiveness of adopting the Treat All policy, regardless of the specific implementation. Therefore, we did not include the actual ART uptake or ART regimen use/change in the modeling.

Missing data occurred in the WHO stage outcome measures. Restricted by the standard software, we used the listwise deletion, i.e., removing missing observations instead of the entire data from the corresponding patient. We applied multiple imputation (MI) for missing data that occurred to the baseline covariates. To enhance the performance of MI, besides the covariates, we added the WHO stage at baseline in the imputation model. We also set the number of replicates to 100 to account for the missing data percentage. Furthermore, we produced the fraction of missing information (FMI), a measure to quantify the loss of information due to missingness while accounting for the amount of information retained by other variables within a data set^22,23^.

All analyses were conducted using R software (version 3.5.1; R Foundation of Statistical Computing, Vienna, Austria). In particular, the *msm* package ^24^ was used to fit multistate models, and the *mice* package was used to impute missing values with the fully conditional specification^25^.

### Multistate Models

Classical survival analysis focuses on the duration from some time origin until the occurrence of a terminal event, a quantity typically denoted by survival or failure time ^26^. If collected, other events before the terminal event may provide more detailed information on the process, enabling more precision in the characterization of disease progression and allowing for the assessment of prognostic factors that may influence the progression. Multistate models (MSMs) provide a framework for analyzing such event history data by modeling the probability of evolving from one state to another over time. Given the presence of four distinct WHO clinical stages with death as the absorbing state in the CA-IeDEA data, MSM is a natural choice to model HIV disease progression.

When a patient demonstrates at least one clinical condition among a WHO stage criterion at a visit, the patient is irreversibly assigned to the most advanced stage in the medical record thereafter^27^. Therefore, we aligned our analysis to this coding standard to define allowable transitions, excluding reverse transitions to preserve the irreversible property. Figure 2 shows the structural transitions with the CA-IeDEA data. The admitted transitions were uni-directional across the WHO stage 1, 2, 3, and 4 and from any such stage to death.

**Figure 2.**
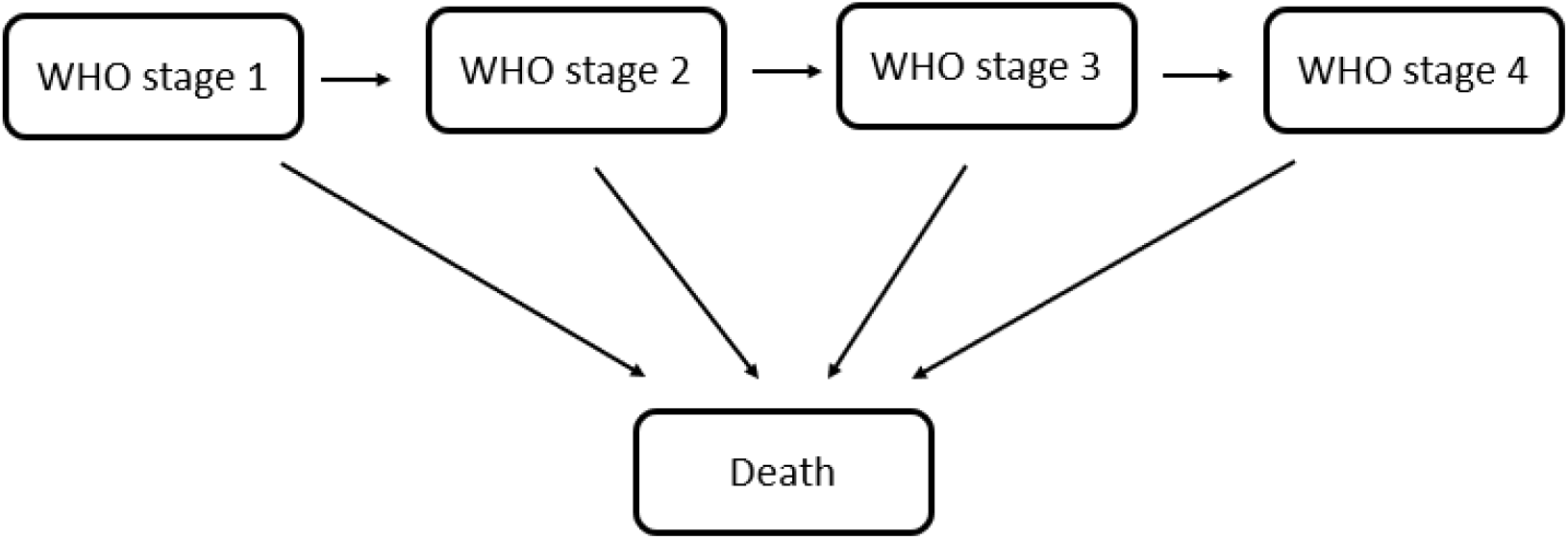
Structural transitions admitted for the HIV disease progression

For this analysis, we employed the so-called continuous-time multistate models. The continuous-time model allows the transition between states to occur at unknown times within the observed time points, including multiple transitions. The model uses all data available (i.e., from patients with two or more observations, including censoring time) to estimate the transition intensity (also known as *transition hazard*) between states. For a detailed introduction to multistate models, we refer to standard references ^28-31^. Here, we give a brief description to allow a definition of the quantities relevant for this paper. Let ***X***_***t***_ denote the state an individual is in at time *t*. Events are then modeled as transitions between the states, so a progression from the asymptomatic to mild symptom stage would be modeled as a 1 → 2 transition. A critical quantity in multistate modeling is the transition hazard, ***λ***_***ij***_(***t***), that is, the *instantaneous risk* to move from state *i* to *j* at time *t*, defined as

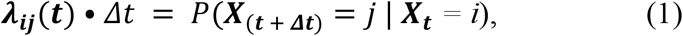

where *P*(***X***_(***t*** + ***Δt***)_ **= *j*** | ***X***_***t***_ *= i*) is the transition probability moving from state *i* to *j* at time *t* (or, more precisely, when *Δt* approaches zero). The transition hazard can be modeled as

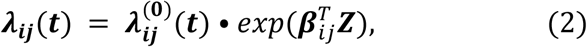

Where 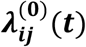 is the baseline hazard function, 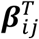 is the transition-specific regression coefficient (i.e., the log hazard ratio), and ***Z*** is the vector of independent variables and covariates (e.g., Treat All treatment, age, gender, etc.). For the baseline hazard function, we used an exponential distribution as it permitted the modeling of the HIV disease process from time zero (even if certain patients had already progressed into a non-normal stage of disease at the enrollment) owing to the memoryless property of the distribution ^32^. Note that when there are only two states and the baseline hazard function is unspecified, model (2) reduces to the commonly used Cox proportional hazard model.

## Results

We had 9,778 patients whose enrollment date in HIV care was within the country’s respective TAG or NUG cohort window. We excluded 732 (7.4%) patients without WHO stage at baseline. Therefore, 9,046 patients (4,607 in the TAG cohort and 4,439 in the NUG cohort) were eligible for the target trial and included in the analysis. Over the entire sample, the median/mean/3^rd^ quantile on the frequency of observations (including time zero, intermittent visits, transferring out, death, and end of cohort timepoints) per person was 2.0/4.4/4.0 and 2.0/4.5/4.0 for the TAG and NUG cohort, respectively. The total accumulated person-years were 5,006 and 4,409 for the TAG and NUG cohort, respectively.

Table 1 describes the enrollment in the target trial from each country. It also summarizes the distribution of the outcome and other covariates at baseline. The people in the NUG cohort were sicker in general, where the percentages of mild/advanced/severe conditions were slightly higher than people in those conditions from the TAG cohort. The imbalance warranted the necessity of covariates adjustment. The mean age at baseline for the patients in the TAG cohort was 33.5 (SD = 12.9) years and 32.0 (SD = 13.4) years for the patients in the NUG cohort. Both cohorts were majority female, comprising 59.2% of the TAG cohort and 62.2% of the NUG cohort. A slightly more proportion of patients were admitted during the dry season (December to April) in the TAG cohort (53.1%) than those in the NUG cohort (48.4%). The median CD4 count at baseline was 378 (IQR = 207-560) for the TAG cohort and 399 (IQR = 239-605) for the NUG cohort in the original unit of cells in a cubic millimeter (cells/mm^3^). Notably, the missing percent for the last two variables were 16% for marital status and 74% for CD4 count, which was high.

**Table 1.**
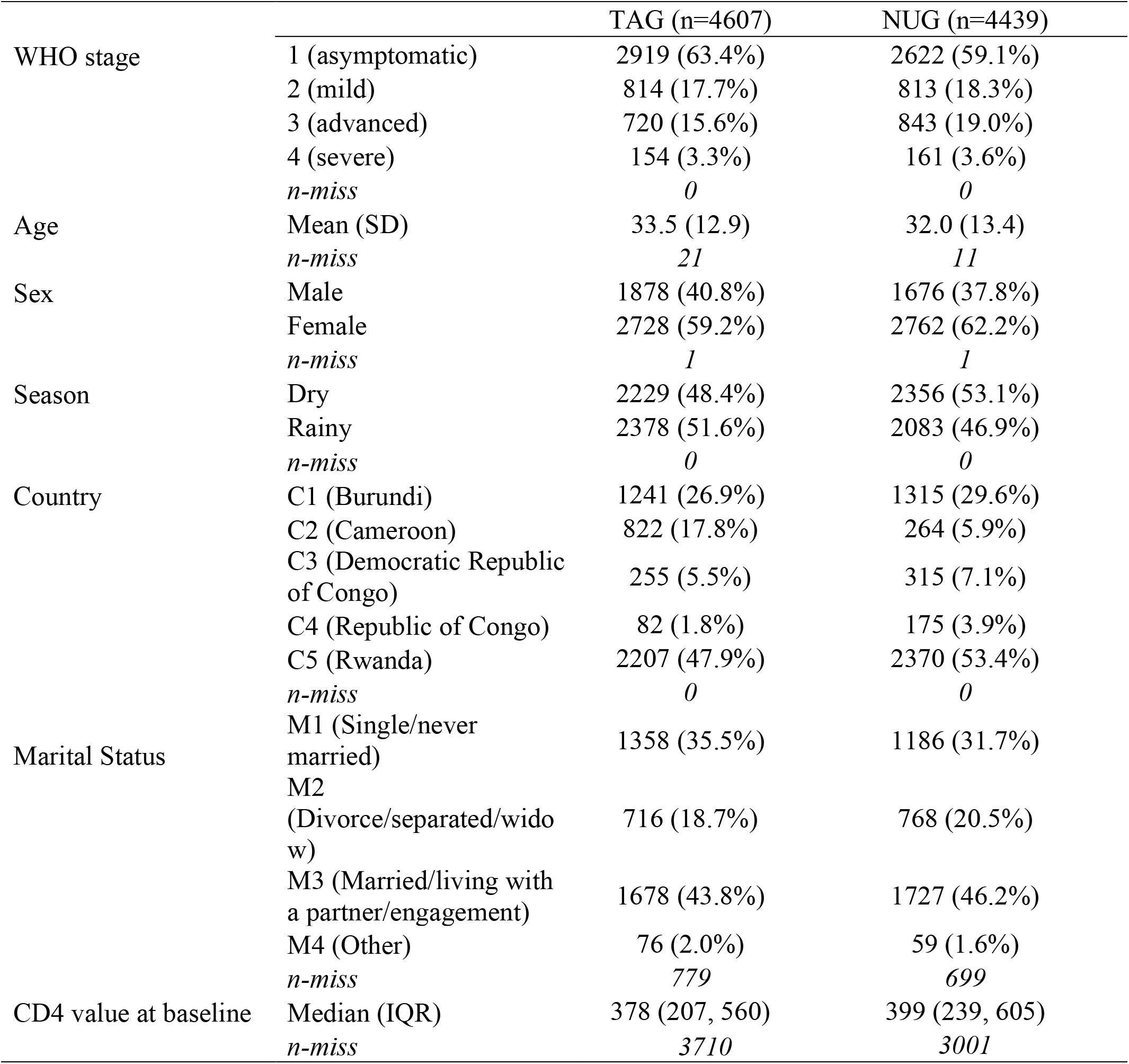
Summary of enrollment by country and patient baseline characteristics

In Table 2, we present the frequency of the observed state-to-state transitions for patients in each cohort. Here we use the term “transition” to describe the observed state status with the understanding that the onset of the transition, when it occurred, can be at an unknown time inside a time interval. There were 15,644 and 15,472 transitions for the TAG and NUG, respectively.

**Table 2.** Observed state-to-state transitions in CA-IeDEA.

|  | TAG |  |  |  |  |  |  | NUG |  |  |  |  |  |  |
| --- | --- | --- | --- | --- | --- | --- | --- | --- | --- | --- | --- | --- | --- | --- |
| From | 1 | 2 | 3 | 4 | 5 | 9 | 99 | 1 | 2 | 3 | 4 | 5 | 9 | 99 |
| 1 | 4667 | 24 | 21 | 7 | 9 | 221 | 2637 | 3459 | 24 | 36 | 2 | 24 | 268 | 2268 |
| 2 | 0 | 2024 | 23 | 3 | 8 | 45 | 759 | 0 | 1784 | 40 | 9 | 14 | 57 | 717 |
| 3 | 0 | 0 | 3578 | 14 | 27 | 27 | 696 | 0 | 0 | 4736 | 16 | 35 | 43 | 825 |
| 4 | 0 | 0 | 0 | 676 | 14 | 7 | 157 | 0 | 0 | 0 | 927 | 17 | 4 | 167 |

1: asymptomatic; 2: mild; 3: advanced; 4: severe; 5: death; 9: transfer out; 99: censored
Note: State transitions that were not admitted (see Figure 2) were assumed to go through the admitted transitions, e.g., $1 \rightarrow 2$ and $2 \rightarrow 3$ for a $1 \rightarrow 3$ transition.

Table 3 presents HRs and aHRs for estimating the effect of Treat All on HIV disease progression (state transition). When adjusting for age, gender, season, and country, patients in the TAG cohort were significantly less likely than NUG cohort patients to transition from WHO stage 1 to stage 2 (aHR = 0.64, 95% CI 0.44 to 0.94) or to transition from WHO stage 1 to death (aHR = 0.37, 95% CI 0.17 to 0.81). There were no statistically significant differences between the TAG and NUG cohorts in the other state transitions. We observed similar results in a sensitivity analysis which included imputed marital status and CD4 count as additional control variables. Table 4 presents the FMI values for each parameter which can be interpreted as the fraction of the total variance (including both between and within imputation variances) that is attributable to between imputation variances, i.e., missingness. We see that imputation did not impact the primary study variable TA (for Treat All), but the impact on CD4 count is in the range of 37-46%. While the point estimates from the sensitivity analysis were valid, caution should be taken to interpret the confidence intervals.

**Table 3.** Results of the multistate model from unadjusted, adjusted, and multiple imputation

| Transition | HR |  | aHR** |  | aHR*** |  |
| --- | --- | --- | --- | --- | --- | --- |
|  | Est | 95% CI | Est | 95% CI | Est | 95% CI |
| 1 → 2 | 0.76 | (0.52,1.10) | 0.64 | (0.44,0.94)* | 0.63 | (0.43,0.93)* |
| 1 → 5 | 0.33 | (0.15,0.73)* | 0.37 | (0.17,0.81)* | 0.35 | (0.16,0.78)* |
| 2 → 3 | 0.64 | (0.45,0.90)* | 0.71 | (0.50,1.01) | 0.72 | (0.51,1.03) |
| 2 → 5 | 0.49 | (0.19,1.28) | 0.58 | (0.18,1.80) | 0.51 | (0.14,1.90) |
| 3 → 4 | 1.09 | (0.63,1.89) | 1.15 | (0.68,1.94) | 1.15 | (0.68,1.95) |
| 3 → 5 | 0.75 | (0.45,1.26) | 0.97 | (0.56,1.66) | 0.95 | (0.54,1.67) |
| 4 → 5 | 0.74 | (0.36,1.51) | 0.91 | (0.44,1.85) | 0.93 | (0.45,1.91) |

Transition coding: 1- asymptomatic; 2 - mild; 3 - advanced; 4 - severe; 5 - death
\*: p-value < 0.05
aHR\*\*: adjusted for age, sex, season, and country
aHR\*\*\*: adjusted for age, sex, season, country, marital, and baseline CD4 counts with multiple imputation (MI)

**Table 4.** FMI from MI for each parameter under each transition with CA-IeDEA

| Transition | Base | TA | Age | Sex | Sea | M2 | M3 | M4 | CD4 | C2 | C3 | C4 | C5 |
| --- | --- | --- | --- | --- | --- | --- | --- | --- | --- | --- | --- | --- | --- |
| 1 → 2 | 0.02 | 0.00 | 0.13 | 0.01 | 0.01 | 0.16 | 0.21 | 0.16 | 0.43 | 0.00 | 0.07 | 0.00 | 0.01 |
| 1 → 5 | 0.01 | 0.00 | 0.04 | 0.02 | 0.00 | 0.10 | 0.08 | 0.09 | 0.42 | 0.01 | 0.00 | 0.00 | 0.02 |
| 2 → 3 | 0.02 | 0.01 | 0.10 | 0.03 | 0.01 | 0.17 | 0.33 | 0.27 | 0.46 | 0.00 | 0.09 | 0.05 | 0.02 |
| 2 → 5 | 0.03 | 0.01 | 0.03 | 0.03 | 0.01 | 0.05 | 0.08 | 0.03 | 0.46 | 0.00 | 0.00 | 0.01 | 0.01 |
| 3 → 4 | 0.01 | 0.01 | 0.10 | 0.01 | 0.00 | 0.11 | 0.21 | 0.24 | 0.38 | 0.01 | 0.04 | 0.00 | 0.03 |
| 3 → 5 | 0.01 | 0.01 | 0.06 | 0.01 | 0.01 | 0.02 | 0.03 | 0.06 | 0.42 | 0.00 | 0.00 | 0.07 | 0.02 |
| 4 → 5 | 0.02 | 0.01 | 0.12 | 0.02 | 0.01 | 0.11 | 0.11 | 0.10 | 0.37 | 0.00 | 0.00 | 0.04 | 0.02 |

Sea: Seasonality; M2: Marital status divorce/separated/widow; M3: marital status Married/living with a partner/engagement; M4: marital status Other; C2: country Cameroon, C3: country Democratic Republic of Congo; C4: country Republic of Congo; C5: country Rwanda.

## Discussion and Conclusion

Our analysis shows that patients enrolling in care after the nationwide adoption of Treat All tended to have fewer transitions to more advanced stages of HIV disease. Notably, significant results were obtained for the asymptomatic to mild and asymptomatic death transition. Moreover, our results indicate that adopting Treat All had no adverse effects on individuals in advanced or severe disease stages. We observed no significant differences in disease progression among these individuals between the TAG and NUG cohorts. Similar findings regarding the no adverse effects for severe people were reported from Boeke et al. ^40^ and Brazier et al. ^32^.

For the observational data where RCT is not feasible, we adopted a target trial in this study, a type of quasi-experiment ^33^. We emulated a hypothetical randomized controlled trial using each country’s pre-Treat All enrollments as the control. With time zero and eligibility defined, we enrolled subjects into the control and treatment cohorts within the same length of the observing period. Subsequently, the designed outcomes (event history of HIV disease in our case) that occurred over the observing window were compared using corresponding statistical models (e.g., multistate regression model in our case). Utilizing the regression models, we conducted the covariates adjustment and sensitivity analysis to control the influence of the confounders. Analytic approach wise, we used intent-to-treat to estimate the overall effectiveness of the Treat All, which is most relevant for evaluating the population impact of policy implementation.

The biological process of HIV disease progression is complex. We approximated it using a continuous-time multistate model that accommodates the interval-censored data where the onset of transition occurred at an unknown time between the observed time points. The most striking feature of multistate models is that they can simultaneously infer the multiple events to estimate the hazard for all desired transitions in the same model. As a comparison, approaches such as those modeling each event separately might result in misleading interpretations and conclusions as they ignore the inter-connection and competing risk among the multiple events.

Other strengths of our study include the use of real-world service delivery data. Pooling the data across 21 clinics from five countries, we had a relatively large sample, enabling exploring the underlying biological process of HIV disease progression. Meanwhile, the relatively rare state advancing transitions occurred within the limited length of follow-up, and the under-reported death prevented us from exploring more biologically plausible models. Those limitations also prevented us from exploring possible heterogeneous effects of Treat All policy implementation across subpopulations, e.g., country and site.

Our study has several other limitations. We used the end of cohort date to censor individuals who were not transferred out and those with no documented death. The censoring was assumed to be non-informative. Loss to follow-up, including unascertained deaths, might be an issue. Among the censored individuals, certain patients could have dropped out or died at a time (unknown from the data) before the censoring time. When the loss to follow-up was informative, e.g., related to the Treat All policy assignment, our estimated hazard ratios can be biased.

Regarding the missing data problem, restricted by the standard software, missingness of survival outcomes was still handled as a complete case analysis, based on an assumption of missing completely at random^34^ that may not be warranted. We attempted a multiple imputation approach for the missing data in the baseline covariates, even though the CD4 count variable had a high missing percentage^35^. The FMI results were still unsatisfactory, suggesting additional auxiliary variables to be collected in practice to account for missing data.

From the target trial design perspective, there might be confounding induced by country-level factors, e.g., the introduction of other policies or economic status changes across the two comparing periods. Our analysis used a categorical country variable to proxy the country-to-country difference. Further identification of detailed country-level factors would be beneficial. In addition, the limited availability of data on patient-level characteristics also restricted our ability to account for demographic variation beyond age, gender and seasonality (and marital status and baseline CD4 in sensitivity analysis) in our MSMs, rendering possible residual confounding.

With the growing and widespread availability of individual-level electronic health record data such as IeDEA, our work represents an approach that takes full advantage of the rich, albeit complex, information about longitudinal individual-level outcomes in large, real-world datasets. Our findings regarding the impact of the national Treat All policy implementation on HIV disease progression among people living with HIV can be informative for programmatic policy implementation, impact evaluation, and research purposes.

## Data Availability

All data produced in the present study are available upon reasonable request to the authors

## Acknowledgements

The project is partially supported by a pilot grant from Einstein-Rockefeller-CUNY (ERC-CFAR) Center for AIDS Research. Central Africa IeDEA is supported by the National Institutes of ‘Health’s National Institute of Allergy and Infectious Diseases (NIAID), the *Eunice Kennedy Shriver* National Institute of Child Health & Human Development (NICHD), the National Cancer Institute (NCI), the National Institute on Drug Abuse (NIDA), the National Heart, Lung, and Blood Institute (NHLBI), the National Institute on Alcohol Abuse and Alcoholism (NIAAA), the National Institute of Diabetes and Digestive and Kidney Diseases (NIDDK), the Fogarty International Center (FIC), the National Library of Medicine (NLM), and the Office of the Director (OD) under Award Number U01AI096299 (Central Africa-IeDEA).

